# Muscle mass, strength, power and physical performance and their association with quality of life in older adults, the Study of Muscle, Mobility and Aging (SOMMA)

**DOI:** 10.1101/2023.10.31.23297845

**Authors:** Nora Petnehazy, Haley N. Barnes, Anne B. Newman, Stephen B. Kritchevsky, Steven R. Cummings, Russel T. Hepple, Peggy M. Cawthon

## Abstract

**Background:** Sarcopenia negatively impacts quality of life. It is unclear whether different measures of muscle size, strength, physical performance, and fitness have similar associations with quality of life.

**Objective:** To describe associations of sarcopenia metrics with quality of life outcomes.

**Participants:** Community-dwelling adults aged 70+ years participating in the SOMMA (Study of Muscle, Mobility and Aging) study.

**Design and settings:** Two academic medical centers.

**Measurements:** Measures included muscle size (MRI-muscle volume. D_3_Cr muscle mass); strength and power (grip strength, leg extension power and strength); walking and physical performance (4m and 400m walk, SPPB (Short Physical Performance Battery), stair climb, chair stand); fitness (VO_2_ peak); health related quality of life (EQ-5D); and anthropometrics (weight, height, and body mass index).

Results were stratified by sex. Correlations, scatterplots and linear regression models described the association between various measures of sarcopenia and fitness with overall quality of life score (EQ5D VAS) as a continuous variable. We also quantified differences between sarcopenia and fitness measures by overall QOL (Quality of Life) as a categorical variable (low, medium, high) and by QOL subcomponents (pain and discomfort, problems with usual activities, mobility, anxiety and depression, and problems with self-care) using distributionally appropriate methods.

**Results:** Walking tests and physical performance were most consistently (but modestly) associated with overall quality of life (r∼0.2, p<.001) and its subcomponents. Both men and women several sarcopenia and fitness measures were more strongly associated with pain and usual activity than other QOL components.

**Conclusions:** Poor performance, lower fitness and lower strength are related to worse quality of life, particularly pain, in older adults. Future studies should quantify these relationships longitudinally.

## Introduction

Sarcopenia is an age-related loss of skeletal muscle mass, strength and physical function associated with several harmful outcomes, such as falls, fractures, loss of independence, disability, and even increased mortality rates (1, 27). Sarcopenia is a generalized and progressive muscle disorder that likely negatively impacts an individual’s quality of life, and on the health care system as well (1,1–4). Further, sarcopenia refers to the gradual loss of muscle mass and strength and has been formally recognized as a muscle disease in the International Classification of Disease (ICD-10: M62 (84)) (5). While many studies have been written related to sarcopenia in association with different outcomes, a single diagnostic criterion has not yet been established (5).

Previous reports have investigated the relation between sarcopenia and quality of life (3,4,6–11). One of the papers discussed the relation between sarcopenia and quality of life in a literature review where only 6 studies found to present a connection with different sarcopenia metrics and quality of life (10). Another paper investigated the association between grip strength and life-quality (using SF-36 questionnaire) (4). Other papers discussed specifics such as sarcopenic obesity, sarcopenia in low- and middle-income countries and several cross-sectional associations in specific clinical populations (end-stage liver disease, osteopenia etc.) (3,7–9,11). The association between sarcopenia and quality of life has not been thoroughly explored; specifically, whether different aspects of factors related to sarcopenia (muscle mass, strength, physical performance) have similar relation to overall quality of life.

Therefore, the aim of this study was to describe associations of sarcopenia metrics and quality of life outcomes. We used data collected during SOMMA (Study on Muscle, Mobility and Aging) (12) to evaluate the association between sarcopenia metrics (MRI-muscle volume, grip-strength, 400m walking test, D_3_Cr muscle mass, SPPB test, Stair climb test, Short walk test, Chair stand test, Leg extension power and strength) and health-related quality of life (EQ-5D) in adults aged 70-89 years old participating in SOMMA.

## Methods

### Study cohort and Recruitment

Participants aged 70 and older were recruited for SOMMA from April 2019 to December 2021 at 2 clinical sites, the University of Pittsburgh and Wake Forest University School of Medicine, as described previously (12). The study was approved by WCGIRB and all participants provided written informed consent. Participants completed the SOMMA measures over three days of visit<u>s</u> that comprised the baseline examination. The data that support the findings of this study are available from the Study of Muscle, Mobility and Aging (SOMMA). Data can be obtained via https://sommaonline.ucsf.edu

SOMMA participants must have been able to complete the 400-meter walk; those who appeared as they might not be able to complete the 400m walk at the in-person screening visit completed a short distance walk (4 meters) to ensure their walking speed as ≥ 0.6m/s. Exclusion criteria included body mass index (BMI) > 40 kg/m^2^; individuals were also excluded if they reported an inability to walk one-quarter of a mile or climb a flight of stairs; had an active malignancy. Screenees with contraindications for a muscle biopsy or MRI were also excluded.

Anthropomorphic measures (weight, height) were assessed by balance beam or digital scales (weight) and by wall-mounted stadiometers (height), BMI (body mass index) was calculated as weight (kg)/height (m^2^).

A modified Balke protocol for cardiopulmonary exercise testing was used to measure VO_2_ peak (13). Walking speed was measured over 400 m (14). We also used the Short Physical Performance Battery (SPPB) (15), which includes a timed 4 m walk test, balance test, chair stand test and the expanded SPPB that also includes a short narrow walk test. Leg extension power and strength (one repetition max, 1 repetition max and peak power) were assessed using the Keiser AIR300 or A420 Leg Press system. Grip strength (16) was measured with a Jamar hand-held dynamometer. We measured a stair climbing task, where participants were asked to climb up and down 4 stairs 3 times without stopping to measure functional leg power (17). An MRI scan was taken of the whole body to assess total thigh muscle volume (AMRA Medical). Whole-body D3 Cr muscle mass was measured in participants using a d3 -creatine dilution protocol (18,19). We assessed the level of physical activity by the Community Healthy Activities Model Program for Seniors (CHAMPS) questionnaire to more precisely assess specific types and the context of physical activities at the baseline and annual visits (20,21). We assessed perceived physical and mental fatigability using the Pittsburgh Fatigability Scale (PFS) where higher scores mean greater fatigability (22,23). Clinic interviews and self-administered questionnaires were conducted. Information collected include questions about sex, age, ethnicity, marital status, tobacco (ever smoked current or past), alcohol (drinks/week), number of comorbidities (no osteo or arthritis) based on modified Rochester Epidemiology Project (0, 1, or 2+ comorbidities) (24). We also analyzed the data stratified by sex.

### Quality of life

We measured quality of life by the EQ-5D questionnaire, and the EQ-VAS recorded the respondent’s self-rated health on a vertical visual analogue scale where the endpoints were labelled ‘The best health you can imagine’ and ‘The worst health you can imagine’. This information was used as a quantitative measure of health outcome as judged by the individual respondents (25).

We analyzed the overall quality of life score as a continuous variable and as categories: low: <80, medium: 80-89, and high: 90 to 100.

There are five subcomponents (Mobility, Self-care, Usual activities, Pain & discomfort, Anxiety & depression) of the EQ-5D. These subcomponents each consist of a single question with a three-level answer (generally no problem, some problem, or severe problem/inability). Few participants answered the “worst” condition, so we divided participants into two groups by their answers on EQ-5D questionnaire: with those reporting any problem versus those reporting no problem at all.

### Statistical Approach

Characteristics of the study population stratified by quality-of-life score were compared using chi-square test for categorical variables, ANOVA for normally distributed continuous variables, and a Kruskal-Wallis test for skewed continuous variables. Linear trend across categories (P-trend) was tested for using linear regression for normally distributed continuous variables. P-trend for skewed continuous variables and categorical variables utilized a two-sided Jonckheere-Terpstra test.

Due to the non-normal distribution of variables, we used Spearman correlation coefficients to describe the relationship between each of the sarcopenia and fitness metrics with quality of life. To compare mean values of sarcopenia and fitness measures by the individual QOL components (mobility, self-care, usual activities, pain & discomfort, and anxiety & depression), t-tests and Wilcoxon rank-sum tests were applied for normally distributed and skewed data respectively. These unadjusted means were also stratified by sex. Quality of life components were dichotomized into “no problems” and “at least some problems” categories for this analysis.

## Results

### Participant Characteristics

The characteristics of the participants are reported by sex-specific categories of life-quality based on EQ-5D-VAS questionnaire (0-100) (Table 1.) We found no association between sex, age, race, marital status, and alcohol drinking habits and the level of the reported quality of life categories. In both men and women, greater quality of life category was associated with lower body mass index (BMI), greater activity level (CHAMPS), lower Pittsburgh Fatigability scores (mental and physical), less smoking, and fewer comorbidities (p-trend: <.05).

**Table 1.** Baseline characteristics of participants by Quality of Life categories.

|  | Quality of Life Categories |  |  |  | *P trend* |
| --- | --- | --- | --- | --- | --- |
| | Overall<br>N = 875 | Low<br>Quality<br>of Life<br>($<80$)<br>N = 165 | Medium<br>Quality of<br>Life (80 – 89)<br>N = 300 | High<br>Quality of<br>Life ($\geq 90$)<br>N = 410 | |
| **Characteristics** |  |  |  |  |  |
| Age, years | 76.3 ± 5.0 | 76.8 ± 5.3 | 76.1 ± 4.8 | 76.4 ± 5.0 | 0.635 |
| Women | 519 (59) | 95 (58) | 183 (61) | 241 (59) | 0.974 |
| Non-Hispanic White | 737 (84) | 135 (82) | 249 (83) | 353 (86) | 0.147 |
| Body mass index, kg/m<sup>2</sup> | 27.6 ± 4.6 | 28.4 ± 4.8 | 28.0 ± 4.6 | 27.0 ± 4.4 | $<.001$ |
| Married/Married-Like Relationship | 444 (51) | 73 (44) | 154 (51) | 217 (53) | 0.093 |
| Ever smoked (current or past) | 381 (44) | 80 (49) | 138 (46) | 163 (40) | 0.027 |
| Alcohol intake, drinks/wk | 2.8 ± 4.5 | 3.1 ± 5.5 | 2.5 ± 4.4 | 2.8 ± 4.1 | 0.340 |
| CHAMPS: Caloric expenditure/week in all exercise-related activities | 3762.2 ± 3051.5 | 2747.2 ± 2439.3 | 3772.4 ± 2694.8 | 4163.3 ± 3409.8 | $<.001$ |
| Pittsburgh Mental Fatigability Score, (0-50) | 7.7 ± 7.7 | 10.8 ± 9.2 | 8.8 ± 7.9 | 5.7 ± 6.2 | $<.001$ |
| Pittsburgh Physical Fatigability Score, (0-50) | 15.8 ± 8.7 | 20.7 ± 8.6 | 17.1 ± 8.3 | 12.9 ± 7.8 | $<.001$ |
| Multimorbidity | | | | | $<.001$ |
| 0 Chronic conditions | 371 (43) | 46 (29) | 113 (38) | 212 (52) |  |
| 1 Chronic condition | 342 (40) | 56 (35) | 130 (44) | 156 (38) |  |
| 2+ Chronic conditions | 152 (18) | 58 (36) | 54 (18) | 40 (10) |  |

Data shown as n (%), mean ± SD
P-trend for normally distributed continuous variables are from linear regression.
P-trend for skewed continuous variables and categorical variables are from a two-sided Jonckheere-Terpstra Test for trend.

We also studied the correlation between the overall quality of life scores, based on the visual analogue scale (VAS) included in EQ-5D; (self-rated health score: 0-100) versus a handful of sarcopenia measures and VO2 peak test that resulted in a significant Spearman correlation coefficient around +/- 0.20. (Figure 1 and Supplemental Table 1). We found that participants with better perceived health status had better performance on the 400 m walk test, 4 m walk test, chair stand test, stair climb test and on the VO_2_ peak test; other sarcopenia measures had weaker correlations with quality of life.

**Figure 1.**
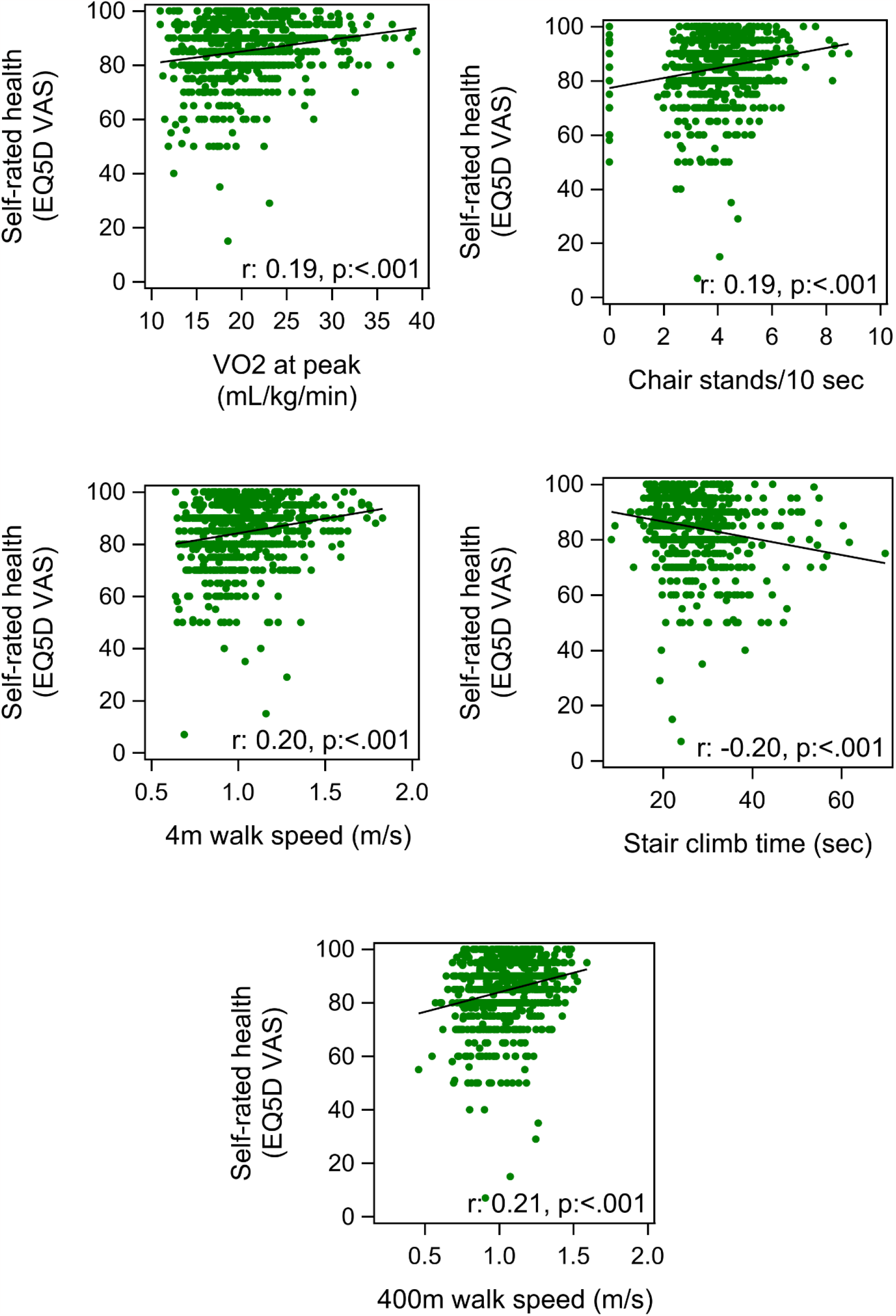
Scatterplots for EQ-5D VAS vs the 5 of x sarcopenia measures having a significant Spearman correlation coefficient around +/- 0.20.

### Mobility QOL

We found significant differences in muscle and performance metrics by the EQ-5D mobility measure (Supplemental Tables 2. and 3.). Men and women with low self-reported mobility had lower D_3_Cr muscle mass (p-value ≤ .006), while total thigh fat free muscle volume (measured with MRI) was not significantly associated with low self-reported mobility. Men and women with low mobility also had worse performance on the stair climb test, chair stand test (10sec) and lower leg power (p-value ≤ 0.002). However, leg strength (1 RM) was significantly associated with low self-reported mobility in women but not in men. Grip strength was not significantly associated with low self-reported mobility in men or women.

Both men and women with low self-reported mobility had significantly worse performance the following tests: SPPB, 400m walk speed, 4m walk speed, narrow walk speed and cardiorespiratory fitness, measured with VO_2_ peak oxygen consumption (ml/kg/min) (p-value ≤ 0.005 for all).

### Self-care QOL (showering, dressing etc.)

10 participants reported any problem with self-care, therefore we do not report comparisons based on this subcategory of quality of life.

### Usual activities QOL (e.g. work, study, housework, family and leisure activities)

Women with lower self-rated function in usual activities had lower muscle mass (D_3_Cr muscle mass and total thigh fat free muscle volume (MRI)); lower grip strength; and lower leg power and strength; worse performance on stair climb test, chair stand test, SPPB, VO_2_ peak test (cardiorespiratory fitness), and on walking tests (400 m, 4 m, narrow) (Supplemental Table 7.).

Men with lower self-reported function in usual activities had lower muscle mass (D_3_Cr muscle mass), lower performance on stair climb test, SPPB, VO_2_ peak (cardiorespiratory fitness) and walking tests (400 m, 4 m, narrow) (p-value ≤ 0.051) (Supplemental Table 6.). In men we found no significant association between lower self-rated function in usual activities and total thigh fat free muscle volume (MRI), chair stand test, grip strength, leg strength and leg power.

### Pain and discomfort QOL

Both women and men with greater pain and discomfort had worse values on many tests including stair climb test, chair stand test, grip strength test, leg power, leg strength, SPPB, VO_2_ peak, and walking tests (400 m, 4 m, narrow) (p-value ≤ 0.021) (Supplemental Tables 8. and 9.). In terms of muscle size, pain and discomfort was only associated with fat free muscle volume in women, p-value = 0.037, but not men; pain and discomfort were not related to muscle size in men (Supplemental Tables 8. and 9.).

### Anxiety and depression QOL

In both women and men, we found no significant results in association of self-reported problems of anxiety and depression with muscle mass (D_3_Cr muscle mass and total thigh fat free muscle volume (MRI)), grip strength, leg power, leg strength, and narrow walk test (p-value ≥0.070) (Supplemental Tables 10. and 11.). In both sexes, we found that participants with higher level of depression and anxiety have lower performance on stair climb and chair stand tests (p-value ≤ 0.058). In men participants who have higher level of anxiety and depression have poorer performance on SPPB test, VO_2_peak (cardiorespiratory fitness) and on the 4 m walk test (p-value ≤ 0.021); these associations were not significant in women. Women with higher self-reported anxiety and depression had lower performance on the 400 m walk test (p-value <= 0.022) whereas in men there was no significance observed (Supplemental Tables 10. and 11.).

## Discussion

Generally, we found that more favorable levels of muscle mass, strength and physical performance were associated with better self-reported quality of life in both men and women. The study also shows clear positive associations between our observed sarcopenia metrics and the EQ-5D subcomponents (Mobility, Self-care, Usual activities, Pain & discomfort, Anxiety & depression).

We found similarities as well as differences in the relation between quality of life and sarcopenia metric between the two sexes. Both in men and women we found that self-reported mobility was strongly associated with most of our sarcopenia metrics (except leg strength and grip strength in men, and muscle mass (MRI) in women). In both men and women, the EQ-5D subcomponent, pain & discomfort, was strongly associated with the majority of our sarcopenia metrics, except for muscle mass measures. For both men and women, we found the fewest connections between self-care and the sarcopenia metrics we used. We speculate that this is because the men and women in SOMMA were generally healthy with relatively few problems with self-care. In women we found that, anxiety & depression problems were associated with only a few of the sarcopenia metrics (only associations were seen for 400 m walk speed, chair stand, and stair climb). In men, while we found more sarcopenia metrics significantly associated with anxiety & depression, it was still the subcomponent with the least connections with the studied sarcopenia metrics. The usual activities subcomponent had the most consistent association with all our observed metrics in case of women and nearly all in men (all except leg power, leg strength, grip strength and muscle mass (D_3_Cr)).

For both men and women we found that some of these sarcopenia metrics were consistently associated with three or more EQ-5D subcomponents: 400 m walk, 4 m walk, narrow walk, VO_2_ peak, stair climb, and chair stand tests. Muscle mass measures (MRI, D_3_Cr) were measures that were the least consistently associated with the EQ-5D subcomponents.

Grip strength, a measure that is routinely included in consensus sarcopenia definitions, surprisingly showed no association with many of the EQ-5D subcomponents such as self-rated mobility, usual activities, pain & discomfort and self-care in men. In women, the pattern was slightly different as grip strength was significantly associated with some of the EQ-5D subcomponents: self-rated mobility, usual activities, and pain & discomfort. The results suggest that we might reconsider the potential impact of grip strength on global quality of life.

On the other hand, walking tests (400 m, 4 m, narrow walk, VO_2_ peak) were strongly related to quality of life in older adults. We consider the VO_2_peak measure a walking test as it was completed on a treadmill. Participants with better cardiopulmonary fitness had significantly higher score on all EQ-5D subgroups in case of men, and on most of them in women. Our study suggests that walking programs, regular physical exercise to maintain fitness, maintain better quality of life as well as a hypothesis to be tested in appropriately studies (13,26,27). We found similar association of chair stand test and stair climb tests to quality of life in line with previous studies (1,5,28).

SOMMA has many strengths. It is a well characterized cohort with a unique constellation of assessments of muscle mass, muscle strength and muscle performance. However, the SOMMA includes mostly non-Hispanic White participants. Further, the population was selected to have no contraindications for MRI (e.g., implanted metal) and biopsy (e.g., no use of blood thinners) and was nondisabled, and so the generalizability of these associations should be explored in other populations We used a global measure of quality of life, rather than a sarcopenia-specific measure of quality of life. We are aware of Sarc-F and Sar-QOL questionnaires and we did not use those in SOMMA as we have more precise physical performance and strength measures than the Sarc-F self-rated questions.

We also covered the main dimensions of Sar-QOL in EQ-5D questionnaire, which is enough to measure quality of life factors broadly for our research aim.

Many SOMMA participants rated their quality of life as high or very high; the figures demonstrate this ceiling effect. This may have limited our ability to determine associations between sarcopenia measures and quality of life, particularly for distinguishing amongst high and very high quality of life rating.

In this observational study we used unadjusted tables as we wanted to first examine the level of associations to determine the simple relation between our measures of strength, power, muscle size, and fitness with quality of life. Based on our findings, further work should endeavor to determine whether these associations are casual, for example, by measuring quality of life as an outcome in clinical trials that aim to increase muscle size or strength.

Overall, results of our study show that poor performance and strength are related to worse quality of life in older adults. Generally, we found that measures of walking ability and physical performance were most consistently associated with overall quality of life and its subcomponents. Finally, we found that pain (in men and women) and problems with usual activities (in women) were most consistently associated with a wide variety of strength and performance measures. Future tudies should investigate whether changes in these measures of sarcopenia are related to declines in quality of life.

## Data Availability

All data produced in the present study are available upon reasonable request to the authors.

## Funding

The Study of Muscle, Mobility and Aging is supported by funding from the National Institute on Aging, grant number AG059416.

Study infrastructure support was funded in part by NIA Claude D. Pepper Older American Independence Centers at University of Pittsburgh (P30AG024827) and Wake Forest University (P30AG021332) and the Clinical and Translational Science Institutes, funded by the National Center for Advancing Translational Science, at Wake Forest University (UL1 0TR001420).

## Conflict of Interest

S.R.C. and P.M.Ca. are consultants to Bioage Labs. All other authors declare no conflict of interest.

## Acknowledgements

We acknowledge all the staff and investigators listed (12), and we thank all the SOMMA participants who enabled this research.

Nora Petnehazy gratefully acknowledges financial support for this publication by the Fulbright Program, which is sponsored by the U.S. Department of State and the Hungarian American Fulbright Commission. Nora Petnehazy also acknowledges financial support by Rosztoczy Foundation.

## Supplemental material

**Supplemental Table 1.** Spearman Correlations for sarcopenia and fitness measures with the EQ-5D VAS.

| Characteristic | N | Spearman<br>Correlation<br>Coefficient | *P-value* |
| --- | --- | --- | --- |
| D<sub>3</sub>Cr muscle mass (kg) | 824 | 0.02 | 0.532 |
| D<sub>3</sub>Cr muscle mass/wgt | 824 | 0.12 | <.001 |
| Total thigh muscle volume (L) | 824 | 0.04 | 0.272 |
| Stair climb time (seconds) | 843 | -0.20 | <.001 |
| Stair climb average functional power (Watts) | 836 | 0.06 | 0.061 |
| Stairs ascended per second (stairs/sec) | 848 | 0.16 | <.001 |
| Chair Stands per 10 sec (chair stands/10 sec) | 874 | 0.19 | <.001 |
| SPPB score (0-12) | 874 | 0.20 | <.001 |
| Walk speed from 400m walk (m/s) | 875 | 0.21 | <.001 |
| Total 400m walk time (sec) | 875 | -0.21 | <.001 |
| VO<sub>2</sub>peak (mL/kg/min) | 816 | 0.19 | <.001 |
| VO<sub>2</sub>peak (without weight adjustment) (mL/min) | 816 | 0.06 | 0.090 |
| 4m walk speed (m/s) | 875 | 0.20 | <.001 |
| Narrow walk speed in m/s using best time(calc) | 798 | 0.14 | <.001 |
| Maximum grip strength (kg) | 869 | 0.04 | 0.203 |
| RPE at 1-RM | 840 | 0.01 | 0.768 |
| Leg Strength: 1 Repetition Maximum | 840 | 0.09 | 0.008 |
| Highest peak power across 40-70% of 1RM (*Watts*) | 840 | 0.06 | 0.090 |
| Highest peak power across 40-70% of 1RM<br>standardized to weight (*Watts/wgt*) | 840 | 0.13 | <.001 |

**Supplemental Table 2.** Sex specific association between the observed sarcopenia metrics, VO2 at peak and an EQ-5D subcomponent, Mobility in men.

| Characteristic | No problem in<br>“walking<br>about”<br>N = 300 | At least some<br>problem in “walking<br>about”<br>N = 56 | *P-value* |
| --- | --- | --- | --- |
| D<sub>3</sub>Cr muscle mass/wgt | 0.32 ± 0.07 | 0.30 ± 0.06 | 0.006 |
| Total thigh muscle volume (L) | 11.25 ± 1.62 | 11.11 ± 1.55 | 0.576 |
| Stair climb time (seconds) | 24.87 ± 5.58 | 29.34 ± 7.89 | <.001 |
| Chair Stands per 10 sec (chair stands/10 sec) | 4.12 ± 1.20 | 3.32 ± 1.67 | <.001 |
| SPPB score (0-12) | 10.48 ± 1.53 | 9.14 ± 2.24 | <.001 |
| Walk speed from 400m walk (m/s) | 1.11 ± 0.16 | 0.95 ± 0.18 | <.001 |
| VO<sub>2</sub>peak (mL/kg/min) | 22.73 ± 4.92 | 19.80 ± 4.55 | <.001 |
| 4m walk speed (m/s) | 1.08 ± 0.19 | 0.97 ± 0.20 | <.001 |
| Narrow walk speed (m/s) | 1.06 ± 0.23 | 0.95 ± 0.26 | 0.005 |
| Maximum grip strength (kg) | 36.99 ± 8.08 | 37.30 ± 8.12 | 0.790 |
| Leg Strength: 1 Repetition Maximum | 219.68 ± 59.79 | 203.73 ± 53.14 | 0.122 |
| Highest peak power across 40-70% of 1RM<br>standardized to weight (*Watts/wgt*) | 5.83 ± 1.78 | 4.99 ± 1.72 | 0.002 |

Data shown as mean ± SD
P-values for continuous variables from a t-test for normally distributed data, a Wilcoxon rank-sum test for skewed data.

**Supplemental Table 3.** Sex specific association between the observed sarcopenia metrics, VO2 at peak and an EQ-5D subcomponent, Mobility in women.

| Characteristic | No problem in<br>“walking<br>about”<br>N = 445 | At least some<br>problem in “walking<br>about”<br>N = 74 | *P-value* |
| --- | --- | --- | --- |
| D<sub>3</sub>Cr muscle mass/wgt | 0.27 ± 0.06 | 0.25 ± 0.06 | 0.001 |
| Total thigh muscle volume (L) | 7.48 ± 1.09 | 7.23 ± 1.11 | 0.094 |
| Stair climb time (seconds) | 26.40 ± 6.86 | 32.74 ± 10.79 | <.001 |
| Chair Stands per 10 sec (chair stands/10 sec) | 4.03 ± 1.15 | 3.32 ± 1.25 | <.001 |
| SPPB score (0-12) | 10.30 ± 1.60 | 9.18 ± 2.03 | <.001 |
| Walk speed from 400m walk (m/s) | 1.04 ± 0.17 | 0.91 ± 0.18 | <.001 |
| VO<sub>2</sub>peak (mL/kg/min) | 18.98 ± 4.10 | 17.06 ± 3.88 | <.001 |
| 4m walk speed (m/s) | 1.03 ± 0.20 | 0.94 ± 0.20 | <.001 |
| Narrow walk speed (m/s) | 0.98 ± 0.23 | 0.85 ± 0.22 | <.001 |
| Maximum grip strength (kg) | 23.41 ± 5.45 | 22.14 ± 6.17 | 0.070 |
| Leg Strength: 1 Repetition Maximum | 142.51 ± 37.81 | 131.02 ± 32.72 | 0.035 |
| Highest peak power across 40-70% of 1RM<br>standardized to weight (*Watts/wgt*) | 4.02 ± 1.17 | 3.34 ± 1.16 | <.001 |

Data shown as mean ± SD
P-values for continuous variables from a t-test for normally distributed data, a Wilcoxon rank-sum test for skewed data.

**Supplemental Table 4.** Sex specific association between the observed sarcopenia metrics, VO2 at peak and an EQ-5D subcomponent, Self-care in men.

| Characteristic | No problem with self-care (showering, dressing, etc.)<br>N = 352 | At least some problem with self-care (showering, dressing, etc.)<br>N = 4 | P-value |
| --- | --- | --- | --- |
| D<sub>3</sub>Cr muscle mass/wgt | 0.32 ± 0.07 | 0.26 ± 0.05 | 0.109 |
| Total thigh muscle volume (L) | 11.24 ± 1.61 | 10.41 ± 1.21 | 0.257 |
| Stair climb time (seconds) | 25.47 ± 6.14 | 33.93 ± 6.92 | 0.033 |
| Chair Stands per 10 sec (chair stands/10 sec) | 4.01 ± 1.30 | 3.18 ± 2.19 | 0.214 |
| SPPB score (0-12) | 10.29 ± 1.71 | 8.75 ± 3.20 | 0.242 |
| Walk speed from 400m walk (m/s) | 1.09 ± 0.17 | 0.84 ± 0.15 | 0.004 |
| VO<sub>2</sub>peak (mL/kg/min) | 22.32 ± 4.97 | 16.65 ± 0.35 | 0.053 |
| 4m walk speed (m/s) | 1.07 ± 0.20 | 0.81 ± 0.10 | 0.007 |
| Narrow walk speed (m/s) | 1.05 ± 0.23 | 0.79 ± 0.26 | 0.061 |
| Maximum grip strength (kg) | 37.08 ± 8.06 | 33.50 ± 9.15 | 0.379 |
| Leg Strength: 1 Repetition Maximum | 217.20 ± 59.07 | 225.00 ± 66.14 | 0.703 |
| Highest peak power across 40-70% of 1RM standardized to weight (Watts/wgt) | 5.71 ± 1.79 | 5.28 ± 2.00 | 0.707 |

Data shown as mean ± SD
P-values for continuous variables from a t-test for normally distributed data, a Wilcoxon rank-sum test for skewed data.

**Supplemental Table 5.** Sex specific association between the observed sarcopenia metrics, VO2 at peak and an EQ-5D subcomponent, Self-care in women.

| Characteristic | No problem with self-care (showering, dressing, etc.)<br>N = 514 | At least some problem with self-care (showering, dressing, etc.)<br>N = 5 | P-value |
| --- | --- | --- | --- |
| D<sub>3</sub>Cr muscle mass/wgt | 0.27 ± 0.06 | 0.23 ± 0.05 | 0.180 |
| Total thigh muscle volume (L) | 7.45 ± 1.09 | 7.24 ± 2.08 | 0.414 |
| Stair climb time (seconds) | 27.25 ± 7.84 | 31.23 ± 7.90 | 0.179 |
| Chair Stands per 10 sec (chair stands/10 sec) | 3.93 ± 1.19 | 3.94 ± 0.59 | 0.993 |
| SPPB score (0-12) | 10.14 ± 1.72 | 10.00 ± 1.00 | 0.563 |
| Walk speed from 400m walk (m/s) | 1.03 ± 0.18 | 0.89 ± 0.11 | 0.083 |
| VO<sub>2</sub>peak (mL/kg/min) | 18.77 ± 4.12 | 17.77 ± 1.97 | 0.791 |
| 4m walk speed (m/s) | 1.02 ± 0.21 | 0.91 ± 0.07 | 0.207 |
| Narrow walk speed (m/s) | 0.96 ± 0.23 | 0.67 ± 0.31 | 0.011 |
| Maximum grip strength (kg) | 23.28 ± 5.53 | 17.20 ± 7.16 | 0.015 |
| Leg Strength: 1 Repetition Maximum | 140.95 ± 37.32 | 146.40 ± 44.93 | 0.846 |
| Highest peak power across 40-70% of 1RM standardized to weight (Watts/wgt) | 3.93 ± 1.19 | 3.78 ± 1.49 | 0.687 |

Data shown as mean ± SD
P-values for continuous variables from a t-test for normally distributed data, a Wilcoxon rank-sum test for skewed data

**Supplemental Table 6.**
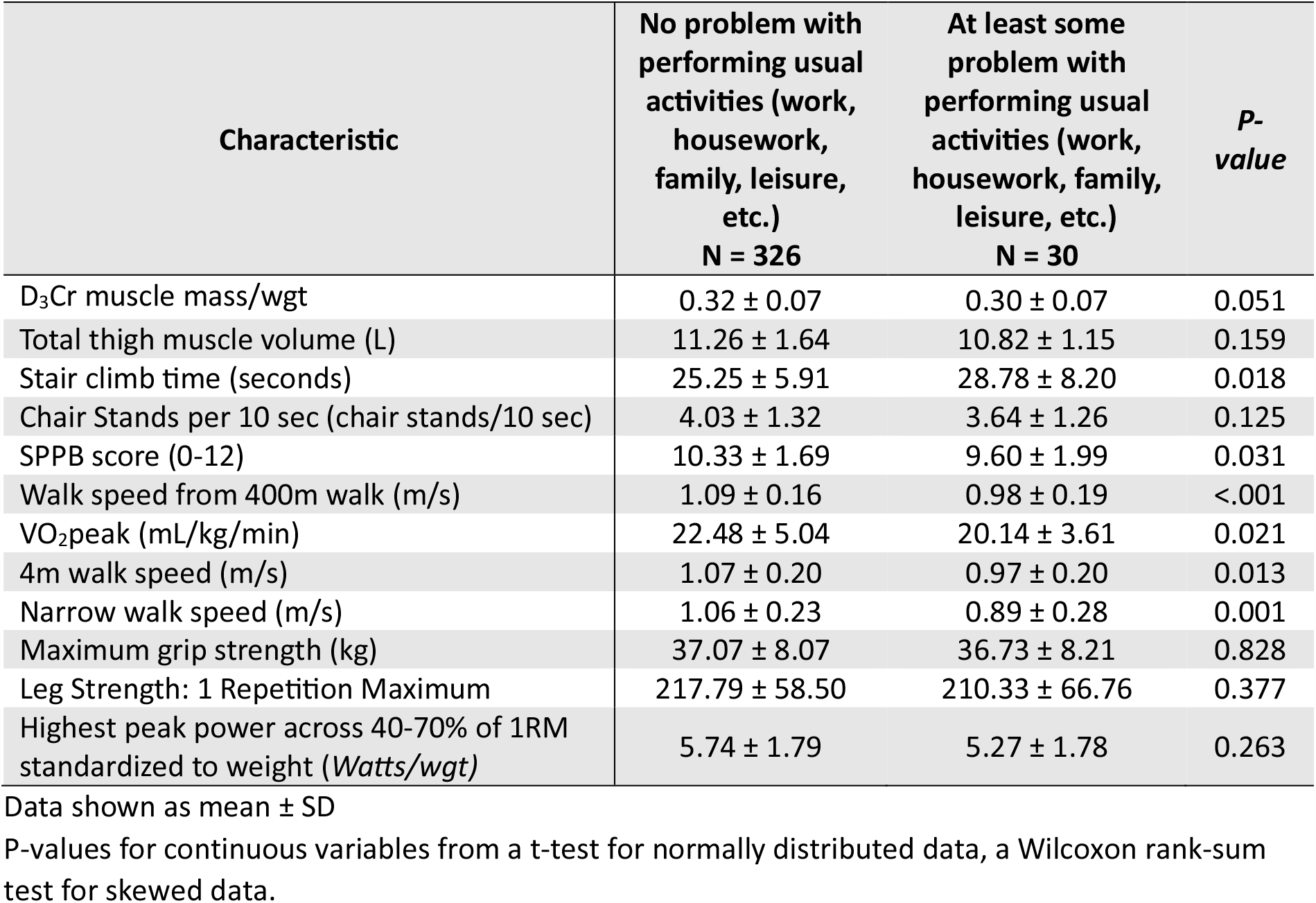
Sex specific association between the observed sarcopenia metrics, VO2 at peak and an EQ-5D subcomponent, Usual activities in men.

**Supplemental Table 7.**
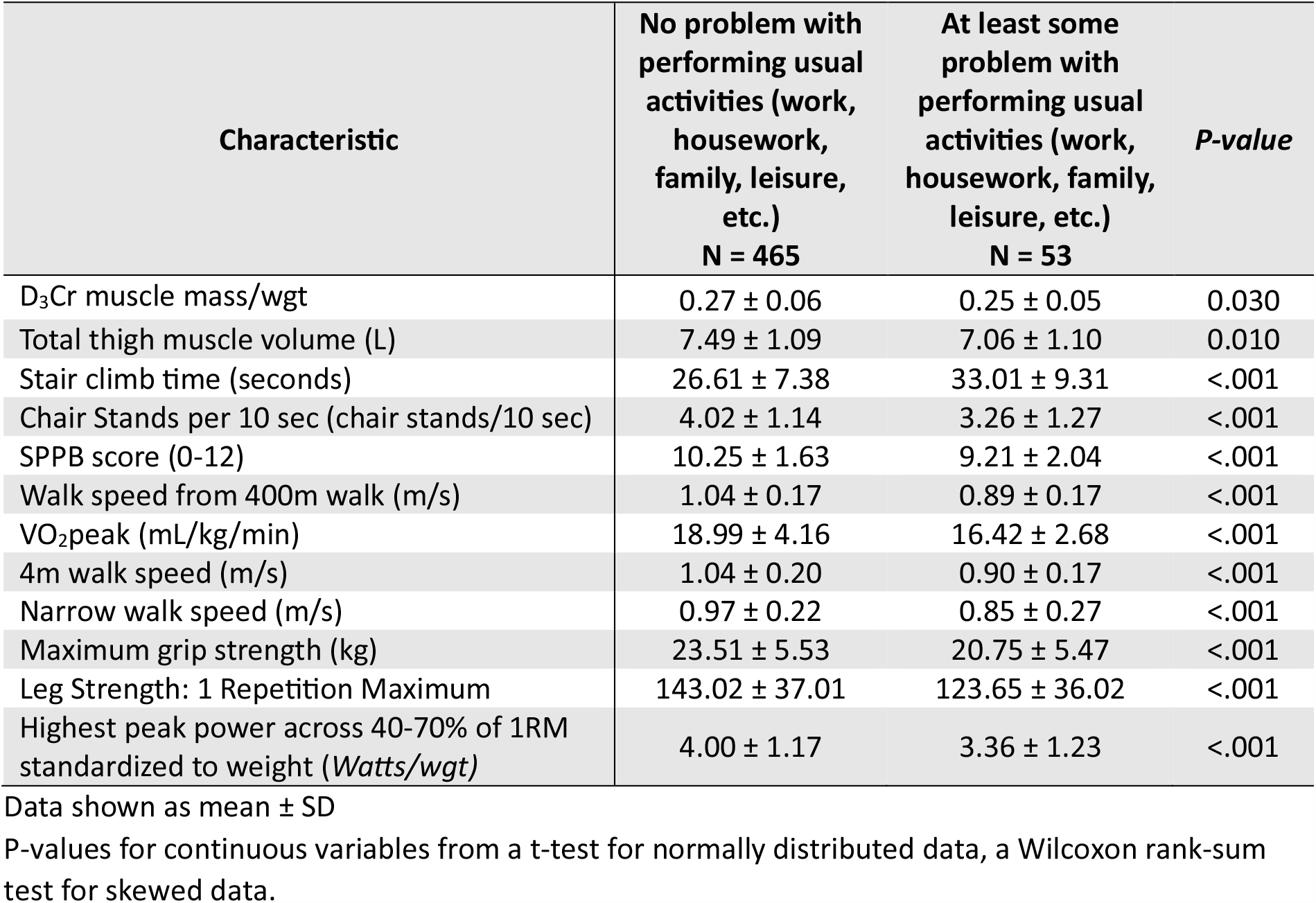
Sex specific association between the observed sarcopenia metrics, VO2 at peak and an EQ-5D subcomponent, Usual activities in women.

**Supplemental Table 8.** Sex specific association between the observed sarcopenia metrics, VO2 at peak and an EQ-5D subcomponent, Pain and discomfort in men.

| Characteristic | No pain or discomfort<br>N = 183 | At least some pain or discomfort<br>N = 172 | P-value |
| --- | --- | --- | --- |
| D<sub>3</sub>Cr muscle mass/wgt | 0.32 ± 0.06 | 0.32 ± 0.07 | 0.159 |
| Total thigh muscle volume (L) | 11.32 ± 1.48 | 11.12 ± 1.74 | 0.176 |
| Stair climb time (seconds) | 24.68 ± 5.76 | 26.48 ± 6.51 | 0.002 |
| Chair Stands per 10 sec (chair stands/10 sec) | 4.25 ± 1.20 | 3.73 ± 1.39 | <.001 |
| SPPB score (0-12) | 10.63 ± 1.45 | 9.89 ± 1.92 | <.001 |
| Walk speed from 400m walk (m/s) | 1.12 ± 0.16 | 1.04 ± 0.17 | <.001 |
| VO<sub>2</sub>peak (mL/kg/min) | 22.92 ± 5.02 | 21.58 ± 4.85 | 0.013 |
| 4m walk speed (m/s) | 1.12 ± 0.20 | 1.01 ± 0.18 | <.001 |
| Narrow walk speed (m/s) | 1.08 ± 0.23 | 1.01 ± 0.23 | 0.006 |
| Maximum grip strength (kg) | 38.08 ± 7.88 | 35.91 ± 8.16 | 0.012 |
| Leg Strength: 1 Repetition Maximum | 224.81 ± 58.09 | 208.87 ± 59.29 | 0.021 |
| Highest peak power across 40-70% of 1RM standardized to weight (Watts/wgt) | 5.96 ± 1.71 | 5.43 ± 1.85 | 0.004 |

Data shown as mean ± SD
P-values for continuous variables from a t-test for normally distributed data, a Wilcoxon rank-sum test for skewed data.

**Supplemental Table 9.** Sex specific association between the observed sarcopenia metrics, VO2 at peak and an EQ-5D subcomponent, Pain and discomfort in women.

| Characteristic | No pain or discomfort<br>N = 257 | At least some pain or discomfort<br>N = 262 | P-value |
| --- | --- | --- | --- |
| D<sub>3</sub>Cr muscle mass/wgt | 0.27 ± 0.06 | 0.26 ± 0.06 | 0.087 |
| Total thigh muscle volume (L) | 7.53 ± 1.03 | 7.36 ± 1.16 | 0.037 |
| Stair climb time (seconds) | 25.67 ± 6.00 | 28.87 ± 9.03 | <.001 |
| Chair Stands per 10 sec (chair stands/10 sec) | 4.10 ± 1.21 | 3.77 ± 1.15 | 0.001 |
| SPPB score (0-12) | 10.39 ± 1.58 | 9.89 ± 1.80 | 0.001 |
| Walk speed from 400m walk (m/s) | 1.06 ± 0.17 | 0.98 ± 0.17 | <.001 |
| VO<sub>2</sub>peak (mL/kg/min) | 19.14 ± 4.10 | 18.35 ± 4.10 | 0.011 |
| 4m walk speed (m/s) | 1.07 ± 0.21 | 0.98 ± 0.19 | <.001 |
| Narrow walk speed (m/s) | 1.00 ± 0.23 | 0.92 ± 0.22 | <.001 |
| Maximum grip strength (kg) | 24.33 ± 5.16 | 22.14 ± 5.75 | <.001 |
| Leg Strength: 1 Repetition Maximum | 145.55 ± 37.64 | 136.31 ± 36.55 | 0.005 |
| Highest peak power across 40-70% of 1RM standardized to weight (Watts/wgt) | 4.19 ± 1.16 | 3.67 ± 1.17 | <.001 |

Data shown as mean ± SD
P-values for continuous variables from a t-test for normally distributed data, a Wilcoxon rank-sum test for skewed data.

**Supplemental Table 10.** Sex specific association between the observed sarcopenia metrics, VO2 at peak and an EQ-5D subcomponent, Anxiety and depression in men.

| Characteristic | Not anxious or depressed<br>N = 322 | At least moderately anxious or depressed<br>N = 34 | P-value |
| --- | --- | --- | --- |
| D<sub>3</sub>Cr muscle mass/wgt | 0.32 ± 0.07 | 0.31 ± 0.05 | 0.447 |
| Total thigh muscle volume (L) | 11.23 ± 1.60 | 11.16 ± 1.76 | 0.766 |
| Stair climb time (seconds) | 25.35 ± 6.13 | 27.37 ± 6.47 | 0.058 |
| Chair Stands per 10 sec (chair stands/10 sec) | 4.05 ± 1.28 | 3.46 ± 1.57 | 0.012 |
| SPPB score (0-12) | 10.36 ± 1.65 | 9.47 ± 2.23 | 0.021 |
| Walk speed from 400m walk (m/s) | 1.09 ± 0.17 | 1.04 ± 0.17 | 0.079 |
| VO<sub>2</sub>peak (mL/kg/min) | 22.50 ± 5.00 | 20.08 ± 4.14 | 0.009 |
| 4m walk speed (m/s) | 1.07 ± 0.20 | 0.99 ± 0.18 | 0.018 |
| Narrow walk speed (m/s) | 1.05 ± 0.24 | 0.98 ± 0.19 | 0.147 |
| Maximum grip strength (kg) | 37.26 ± 7.99 | 34.97 ± 8.64 | 0.116 |
| Leg Strength: 1 Repetition Maximum | 217.77 ± 59.04 | 212.16 ± 59.66 | 0.772 |
| Highest peak power across 40-70% of 1RM standardized to weight (Watts/wgt) | 5.75 ± 1.79 | 5.29 ± 1.79 | 0.241 |

Data shown as mean ± SD
P-values for continuous variables from a t-test for normally distributed data, a Wilcoxon rank-sum test for skewed data.

**Supplemental Table 11.** Sex specific association between the observed sarcopenia metrics, VO2 at peak and an EQ-5D subcomponent, Anxiety and depression in women.

| Characteristic | Not anxious or depressed<br>N = 447 | At least moderately anxious or depressed<br>N = 71 | P-value |
| --- | --- | --- | --- |
| D<sub>3</sub>Cr muscle mass/wgt | 0.27 ± 0.06 | 0.27 ± 0.05 | 0.872 |
| Total thigh muscle volume (L) | 7.46 ± 1.12 | 7.33 ± 0.96 | 0.351 |
| Stair climb time (seconds) | 26.92 ± 7.61 | 29.59 ± 8.87 | 0.004 |
| Chair Stands per 10 sec (chair stands/10 sec) | 3.98 ± 1.18 | 3.65 ± 1.20 | 0.033 |
| SPPB score (0-12) | 10.19 ± 1.69 | 9.83 ± 1.84 | 0.113 |
| Walk speed from 400m walk (m/s) | 1.03 ± 0.18 | 0.98 ± 0.16 | 0.022 |
| VO<sub>2</sub>peak (mL/kg/min) | 18.81 ± 4.19 | 18.31 ± 3.46 | 0.514 |
| 4m walk speed (m/s) | 1.02 ± 0.21 | 1.01 ± 0.20 | 0.782 |
| Narrow walk speed (m/s) | 0.97 ± 0.23 | 0.93 ± 0.23 | 0.211 |
| Maximum grip strength (kg) | 23.31 ± 5.43 | 22.77 ± 6.41 | 0.456 |
| Leg Strength: 1 Repetition Maximum | 141.63 ± 37.77 | 137.18 ± 34.87 | 0.463 |
| Highest peak power across 40-70% of 1RM standardized to weight (Watts/wgt) | 3.97 ± 1.20 | 3.70 ± 1.10 | 0.070 |

Data shown as mean ± SD
P-values for continuous variables from a t-test for normally distributed data, a Wilcoxon rank-sum test for skewed data.

## Notes

### Competing Interest Statement

The authors have declared no competing interest.

### Author Declarations

Ethics committee of the California Pacific Medical Center Research Institute gave ethical approval for this work.

